# FinnDiane LifeOne Study - Impact of ageing on people with type 1 diabetes, a prospective observational cohort study

**DOI:** 10.64898/2026.05.06.26352532

**Authors:** Jenna Nicklén, Susanna Satuli-Autere, Kaarina Rimpeläinen, Anu Dufva, Anni Ylinen, Emilia M.C. Franzén, Marika I. Eriksson, Fanny Jansson Sigfrids, Hanna Öhman, Lena M. Thorn

## Abstract

**Introduction:** Life expectancy for people with type 1 diabetes has increased due to improved treatment of diabetes and its comorbidities, allowing many to reach old age. Still, we lack knowledge of how individuals with type 1 diabetes age. On one hand, those who reach older age can be considered survivors, but on the other hand their long-standing diabetes might still exhibit negative impacts on their health and functional ability. Healthy ageing is the World Health Organization’s priority for this decade. The focus has shifted from chronological age to functional ability, which reflects the ability of individuals to perform meaningful activities. Functional ability is shaped by intrinsic capacity, the environment, and their interaction. Intrinsic capacity encompasses five main domains: cognition, vitality, sensory function, locomotion, and psychological domain. This observational study aims to assess how this vulnerable group of individuals with type 1 diabetes age and to identify factors that contribute to their healthy ageing, intrinsic capacity, and its domains.

**Methods and analysis:** The FinnDiane LifeOne Study is a prospective observational cohort study. We aim to recruit a minimum of 300 individuals with type 1 diabetes from the FinnDiane Study, aged >65, and a minimum of 100 matched controls without insulin-dependent diabetes. The cohort will be comprehensively characterized, including clinical assessment, laboratory tests, questionnaires, and a geriatric assessment of different aspects of functioning ability, with five years intervals. We will compare the individuals with type 1 diabetes to their matched controls. For those with type 1 diabetes, we will further assess which factors from the FinnDiane baseline and trajectories during follow-up predict healthy ageing in above 65-year-olds.

**Ethics and dissemination:** The LifeOne study protocol is approved by the Ethics Committee of HUS Helsinki University Hospital (HUS/4387/2023) and the study adheres to the Declaration of Helsinki. Written informed consent is obtained from each participant. Findings will be published in international peer-reviewed journals with an open access choice.

The study is registered at ClinicalTrials.gov with ID NCT07289204.

**STRENGTHS AND LIMITATIONS OF THE STUDY:**

- This is a prospective observational cohort study with a matched control group.
- For the participants with type 1 diabetes, we have unique and comprehensive longitudinal clinical and genetic data available from approximately participants’ middle age, enabling identification of factors that contribute to their healthy ageing, while accounting for the competing risk of death.
- The cohort is thoroughly characterised regarding diabetes, cardiometabolic health, lifestyle, psychosocial factors, and includes a geriatric assessment, thereby enabling comparison of impact of ageing between individuals with type 1 diabetes and controls without insulin-dependent diabetes.
- The cohort is Caucasian with recruitment from Southern Finland, potentially limiting generalisability to other more ethnically diverse populations.

## INTRODUCTION

Ageing with type 1 diabetes is an emerging reality, yet it poses new challenges. Life expectancy among people with type 1 diabetes has improved due to advances in the treatment of both the disease itself and its complications [1]. However, decades of living with type 1 diabetes and chronic dysglycaemia inevitably has adverse health effects, leading to an increased burden of micro- and macrovascular disease as well as higher mortality rates [2, 3]. With scarce research on older adults with type 1 diabetes, ageing in type 1 diabetes has been identified as a significant research gap [4].

Diabetes is associated with accelerated aging [4]. Type 1 diabetes-specific consequences include accentuated vascular ageing that occurs 15–20 years earlier than in the general population [5]. Despite an increased risk of micro- and macrovascular complications with longer duration of type 1 diabetes [6, 7], those who survive 50 years with type 1 diabetes tend to have a lower prevalence of diabetic kidney disease, as an indication of survival bias [8]. However, a long disease duration does not necessarily correspond to older age, nor do all elderly individuals have a long disease duration. In studies of older adults with type 1 diabetes, typical characteristics include older diabetes onset age, lower prevalence of smoking, lower mean arterial pressure and HbA1c, lower prevalence of diabetic kidney disease, but still signs of vascular ageing with higher pulse pressure and more cardiovascular disease [10, 11, 12]. Furthermore, with ageing, severe hypoglycaemic episodes are more common and they in turn associate with a greater risk of dementia [13]. Regarding dementia, the incidence is higher in type 1 diabetes compared to the general population, both in younger and older age groups [14]. Interestingly, geriatric syndromes, such as frailty, are common already in rather young individuals with type 1 diabetes and associate with increased mortality and a higher risk of diabetic complications [9]. How frailty relates to the functional ability of individuals with type 1 diabetes is, however, not known.

Healthy ageing is the World Health Organization’s focus of this decade, defined as “the process of developing and maintaining the functional ability that enables wellbeing in older age”, shifting focus from chronological age to functional ability [15]. This functional ability is determined by intrinsic capacity, the environment, and their interaction [15]. The main domains of intrinsic capacity are considered cognition, vitality, sensory, locomotion, and psychological domain [16].

Despite continued efforts to define it, healthy ageing remains a broad and evolving concept lacking a universally accepted definition. Its assessment requires a multidisciplinary approach that accounts for the complexity and diversity of the ageing processes. There are no efforts, so far, to assess healthy ageing, intrinsic capacity and its domains in type 1 diabetes, although the condition is known to cause deficits in several intrinsic capacity domains.

With this study, we aim to deepen our understanding of ageing in people with type 1 diabetes, with a particular focus on factors that contribute to healthy ageing and the preservation of functional ability to promote healthy, functional, and safe ageing in people with type 1 diabetes. We aim to identify the key characteristics of healthy ageing, intrinsic capacity, and its domains in individuals with type 1 diabetes, and to examine how these differ from controls without insulin-deficiency. In addition, we aim to determine which midlife factors in people with type 1 diabetes predict healthy ageing and functional ability in later life.

## METHODS AND ANALYSES

### Study design

The FinnDiane LifeOne Study (ClinicalTrials.gov ID NCT07289204) is an observational cohort study of people >65 years living with type 1 diabetes. The study is a sub-study of the Finnish Diabetic Nephropathy (FinnDiane) Study, a nationwide cohort study with longitudinal follow-up of more than 20 years [17]. The study design of LifeOne Study is prospective but also includes retrospective data from the FinnDiane Study (Figure 1). The LifeOne Study is a single-centre study conducted at the Helsinki University Hospital. We will invite those FinnDiane participants alive and >65 years living in the Helsinki capital region for a comprehensive assessment including a regular FinnDiane study visit and an additional visit specific for this study, consisting of a geriatric assessment. Because the study site is in Helsinki, participants’ area of residence has been limited to a reasonable proximity to Helsinki to avoid placing an undue burden on older participants. The study visits started in January 2024 and we aim to study a minimum of 300 FinnDiane study participants and recruit a minimum of 100 matched controls. The cohort will be prospectively followed every five years, and the relevant registry data will also be collected.

**Figure 1.**
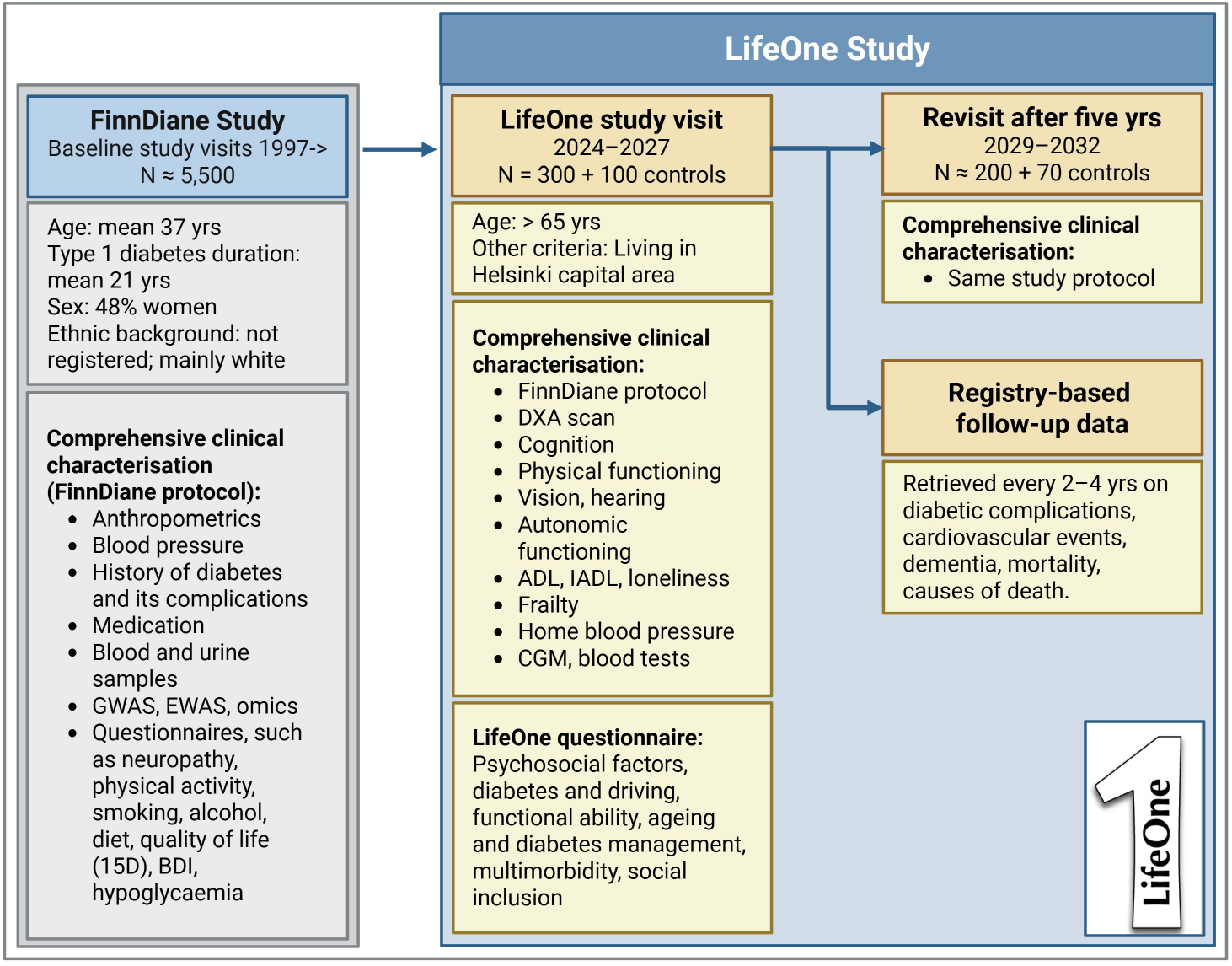
Study design of the LifeOne Study. Created with BioRender.com. GWAS, Genome-Wide Association Study; EWAS, Epigenome-Wide Association Study; BDI, Beck Depression Inventory; DXA, Dual-energy X-ray Absorptiometry; ADL, Activities of Daily Living; IADL, Instrumental Activities of Daily Living; CGM, Continuous Glucose Monitoring.

### Study cohort

#### Participants with type 1 diabetes, >65 years

We will recruit a minimum of 300 participants from the FinnDiane Study for this sub-study. Inclusion criteria include age >65 years and a clinical diagnosis of type 1 diabetes with additional requirement of age at diabetes onset <40 years and insulin treatment initiated within one year of diagnosis.

#### Control participants without insulin-dependent diabetes

We will recruit a minimum of 100 volunteers >65 years, without type 1 diabetes or without other type of diabetes requiring treatment with daily insulin. Controls will be recruited to match the study population for age and sex. As recruitment strategy, we will aim to recruit spouses of the participants to match for socioeconomic factors and lifestyle. For control participants, no retrospective clinical study visits will be available, which means that control participants enable us to do cross-sectional comparisons between individuals with and without type 1 diabetes at the starting point of this sub-study and prospectively during the planned follow-up.

### Sample size calculation

The number of participants with type 1 diabetes is restricted by the number of FinnDiane study participants living in the Helsinki capital region, alive, and aged >65. In 2020, this figure was estimated at 380 potential participants, with a natural increase over upcoming years as the study population ages. Based on these calculations, a target of minimum 300 participants was set. The number of control participants, on the other hand, is restricted by the study visit capacity. A target of minimum 100 control participants will result in enough statistical power to detect between-group differences of medium effect size, but not small effect sizes. To achieve a good power for differences of small effect sizes, both the groups would need to be substantially larger (e.g., 600 for both groups), which is not feasible.

### Regular FinnDiane study visit

The FinnDiane study protocol has been described in detail elsewhere [18].

In brief, the study visit at the FinnDiane research facilities at the Helsinki University Hospital includes the following both for participants with type 1 diabetes and for controls:

- Registration of medical history of diabetes and diabetic complications, as well as current medication, verified from medical records.
- Measurements of body weight, height, waist- and hip circumferences, office blood pressure, and pulse.
- Foot examination of arterial pulses, pressure perception, Achilles tendon reflexes, deformities, and ulcers.
- Cardiovascular status, including ECG, assessment of carotid intima-media thickness by a multiarray echotracking system (Artlab, Esaote, Genova, Italy) and arterial stiffness by applanation tonometry (SphygmoCor, Atcor Medical, Sydney, Australia).
- Body composition and bone mineral density measured with dual-energy X-ray absorptiometry (Lunar iDXA, GE Healthcare, Wauwatosa, WI, US).
- Digital fundus photography (CX-1, Canon Inc., Tokyo, Japan)
- Optical Coherence Tomography Angiography (SOLIX, Optovue Inc., Fremont, CA, US)
- Corneal confocal microscopy (Heidelberg HRT 3 Tomograph, Heidelberg Engineering GmbH, Heidelberg, Germany)
- Blood and urine samples analyzed for standard cardio-kidney-metabolic profile, DNA, multiomics, and stored for future analyses.
- Registration of glucose monitoring from the previous 14 days.
- A large battery of questionnaires including family history, education, working history, lifestyle such as alcohol consumption, diet, food records, smoking, and physical activity based on a Finnish version of the Minnesota leisure time physical activity questionnaire [19, 20]. Further questionnaires include health-related quality of life (15D) [21], Beck Depression Inventory [22], Antonovsky’s sense of coherence [23], neuropathy questionnaires (Michigan Neuropathy Screening Instrument [24], DN4 [25], Survey of Autonomic Symptoms [26]), and history of hypoglycemia.

### LifeOne study visit

The LifeOne study visit is a separate study visit, primarily within a month from the regular FinnDiane study visit, but at least within a year. The geriatric assessment evaluations are performed by a trained study nurse. The study visit includes the following evaluations:

- Cognitive evaluation: Clock-drawing test [27], Animal naming test [28], Mini-Mental State Examination [29], Trail Making Tests A and B [30].
- Physical function: Short Physical Performance Battery [31] .
- Handgrip strength measured by a hydraulic dynamometer (SAEHAN, Saehan Corporation, South Korea).
- Visual accuracy by ESC2000 Illuminated ETDRS Clinical Trial Cabinet (Good-Lite, Eligin, IL, US).
- Audiometry by GSI-18 (Grason Stadler Inc., Eden Prairie, MN, US) after otoscopic examination to ensure clean ear canals.
- Autonomic function by an orthostatic blood pressure test and with a noninvasive, hand-held device (Vagus™, Medicus Engineering ApS, Aarhus, Denmark).
- Registration of Activities of Daily Living (Barthel Index) [32], Instrumental Activities of Daily Living (Lawton scale) [33], Clinical Frailty Scale [34], and perceived loneliness by a validated questionnaire [35].
- Additional laboratory test for complete blood count, thyrotropic hormone, active vitamin B12, folate, and plasma albumin.
- Four days’ home blood pressure measurements (two measurements in the morning and two in the evening in resting state, sitting position) prior to the study visit.
- LifeOne questionnaire assessing living conditions, poverty, retirement age, potential disability pension, diabetes and driving, management of diabetes and challenges with ageing, multimorbidity (list of predefined conditions), frequency of severe hypoglycemia, the Experiences of Social Inclusion Scale [36], and physical functioning (500 m walking) [37].

### Patient and public involvement

Patients and the public were not involved in the design or conduct of the study.

## Data Availability

All data produced in the present work are contained in the manuscript

## DATA ANALYSES PLAN

Data analyses will start at the end of the year 2026 as soon as enough participants and controls have participated. The purpose is to identify the key characteristics of healthy ageing, intrinsic capacity, and its domains in individuals with type 1 diabetes. For this, data-driven approaches such as network analysis will be utilized to assess the interplay within and between the different domains. Furthermore, we will study how the cohort characteristics (e.g., diabetes characteristics, morbidity, poverty, and lifestyle) differ according to level of healthy ageing to understand the interplay between morbidity and functional ability in these older adults with type 1 diabetes. Ageing-related traits (e.g., body composition, sarcopenia, vascular health, hearing loss, and autonomic function) and healthy ageing will then be compared between older adults with type 1 diabetes and the controls without insulin-dependent diabetes. Between-group comparisons will be analysed with appropriate parametric and non-parametric tests, and independent associations determined with multivariable models. In addition, we will determine which midlife factors in people with type 1 diabetes predict healthy ageing later in life, with a special interest in ageing-related biomarkers, such as epigenetic clocks [38], telomere length [39], mitochondrial DNA copy number [40], and polygenic risk scores for ageing-related traits such as muscle strength, longevity, and cognitive function. In these analyses, we will also account for metabolic risk factors, lifestyle, frailty, multimorbidity, and their trajectories during follow-up, as well as account for death as a competing risk.

## ETHICS AND DISSEMINATION

### Ethics

The FinnDiane LifeOne Study protocol was approved by the HUS Regional Committee on Medical Research Ethics (HUS/4387/2023), as was the FinnDiane study protocol (491/E5/2006, 238/13/03/00/2015, and HUS/3313/2018). The study is performed in accordance with the Declaration of Helsinki. Written informed consent is obtained from each participant prior to study participation. If the participant has dementia and at least moderate cognitive impairment, we will ensure written informed consent from the participant’s guardian or next of kin.

The participants have the right to gain information about their results with relevance to their health. All participants will receive a summary of their test results relevant to their health care, including a physician’s interpretation of the results. In case of incidental findings in any testing related to the study, the participants are informed and immediately referred for treatment or medical evaluation.

### Dissemination

Results from the study will be shared via different platforms, such as conference presentations and publications in international peer-reviewed journals with possibility to publish open access. Results will also be disseminated to the public in collaboration with relevant organisations. Individual-level data for the study participants are not publicly available because of consent-related restrictions. After the data collection is finalised, the Readers may propose collaboration to research the individual level data with correspondence with the lead investigator.

## ACKNOWLEDGEMENTS

We gratefully acknowledge all the physicians and nurses at each study centre participating in the collection of the original data for the FinnDiane Study (Online Supplemental material), thus enabling this current sub-study. We thank all participants for their dedication and contribution to this study. We acknowledge the skilled technical assistance of research nurses Anna Sandelin and Kirsi Uljala (Helsinki University Hospital and Folkhälsan Research Center, Helsinki). We thank Anniina Tynjälä for brainstorming the study acronym LifeOne, which highlights both the longevity of individuals with type 1 diabetes (life with type 1) and the importance to cherish the one life we all have (one life).

## AUTHORS’ CONTRIBUTIONS

J.N. and L.M.T. prepared the first draft of the protocol manuscript, while all the authors gave their critical comments and approved the final version of the manuscript. L.M.T. and H.Ö. were head designers of the geriatric assessment; M.I.E., A.Y., and F.J.S. participated in the design of the study visits, and A.D. planned the practicalities for the study visits, including data management. K.R. participated in the data management planning and as the study coordinator oversees the data collection and interprets the test results to the participants. E.M.C.F. participates in quality control of the collected data. S.S.-A. participated in the design of the LifeOne questionnaire. L.M.T. is the principal investigator of this study and takes full responsibility for the content of this work.

