## Supplementary material for "FinnDiane LifeOne Study - Impact of ageing on people with type 1 diabetes, a prospective observational cohort study"

| FinnDiane Study Centers | Physicians and nurses |
| --- | --- |
| Anjalankoski Health Center | S.Koivula, T.Uggeldahl |
| Central Finland Central Hospital, Jyväskylä | T.Forslund, A.Halonen, A.Koistinen, P.Koskiaho, M.Laukkanen, J.Saltevo, M.Tiihonen |
| Central Hospital of Åland Islands, Mariehamn | M.Forsen, H.Granlund, A.-C.Jonsson, B.Nyroos |
| Central Hospital of Kanta-Häme, Hämeenlinna | P.Kinnunen, A.Orvola, T.Salonen, A.Vähänen |
| Central Hospital of Kymenlaakso, Kotka | R.Paldanius, M.Riihelä, L.Ryysy |
| Central Hospital of Länsi-Pohja, Kemi | H.Laukkanen, P.Nyländen, A.Sademies |
| Central Ostrobothnian Hospital District, Kokkola | S.Anderson, B.Asplund, U.Byskata, P.Liedes, M.Kuusela, T.Virkkala |
| City of Espoo Health Center: |  |
| Espoonlahti | A.Nikkola, E.Ritola |
| Tapiola | M.Niska, H.Saarinen |
| Samaria | E.Oukko-Ruponen, T.Virtanen |
| Viherlaakso | A.Lyytinen |
| City of Helsinki Health Center: |  |
| Puistola | H.Kari, T.Simonen |
| Suutarila | A.Kaprio, J.Kärkkäinen, B.Rantaeskola |
| Töölö | P.Kääriäinen, J.Haaga, A-L.Pietiläinen |
| City of Hyvinkää Health Center | S.Klemetti, T.Nyandoto, E.Rontu, S.Satuli-Autere |
| City of Vantaa Health Center: |  |
| Korso | R.Toivonen, H.Virtanen |
| Länsimäki | R.Ahonen, M.Ivaska-Suomela, A.Jauhiainen |
| Martinlaakso | M.Laine, T.Pellonpää, R.Puranen |
| Myyrmäki | A.Airas, J.Laakso, K.Rautavaara |
| Rekola | M.Erola, E.Jatkola |
| Tikkurila | R.Lönnblad, A.Malm, J.Mäkelä, E.Rautamo |
| Heinola Health Center | P.Hentunen, J.Lagerstam |
| Helsinki University Hospital, Department of Medicine, Division of Nephrology | R.Bergdal, T.Claesson, A.Dufva, N.Elonen, M.Eriksson, J.Fagerudd, M.Feodoroff, D.Gordin, P.-H.Groop, O.Heikkilä, K.Hietala, S.Hägg-Holmberg, S.Itkonen, F.Jansson Sigfrids, M.Korolainen, J.Kytö, S.Lindh, J.Nicklén, H.Paajanen, K.Pettersson-Fernholm, K.Rimpeläinen, M.Rosengård-Bärlund, M.Rönnback, L.Salovaara, A.Sandelin, M.Saraheimo, S.Satuli-Autere, R.Simonsen, P.Smidtslund, L.Thorn, H.Tikkanen, J.Tuomikangas, A.Tynjälä, K.Uljala, T.Vesisenaho, J.Wadén, A.Ylinen |
| Herttoniemi Hospital, Helsinki | V.Sipilä |
| Hospital of Lounais-Häme, Forssa | T.Kalliomäki, J.Koskelainen, R.Nikkanen, N.Savolainen, H.Sulonen, E.Valtonen |
| Hyvinkää Hospital | L. Norvio, A.Hämäläinen |
| Iisalmi Hospital | E.Toivanen |
| Jokilaakso Hospital, Jämsä | A.Parta, I.Pirttiniemi |
| Jorvi Hospital, Helsinki University Central Hospital | S.Aranko, S.Ervasti, R.Kauppinen-Mäkelin, A.Kuusisto, T.Leppälä, K.Nikkilä, L.Pekkonen |
| Jyväskylä Health Center, Kyllö | K.Nuorva, M.Tiihonen |
| Kainuu Central Hospital, Kajaani | S.Jokelainen, K.Kananen, M.Karjalainen, P.Kemppainen, A-M.Mankinen, A.Reponen, M.Sankari |
| Kerava Health Center | H.Stuckey, P.Suominen |
| Kirkkonummi Health Center | A.Lappalainen, M.Liimatainen, J.Santaholma |
| Kivelä Hospital, Helsinki | A.Aimolahti, E.Huovinen |
| Koskela Hospital, Helsinki | V.Ilkka, M.Lehtimäki |
| Kotka Health Center | E.Pälikkö-Kontinen, A.Vanhanen |
| Kouvola Health Center | E.Koskinen, T.Siitonen |
| Kuopio University Hospital | E.Huttunen, R.Ikäheimo, P.Karhapää, P.Kekäläinen, M.Laakso, T.Lakka, E.Lampainen, L.Moilanen, S. Tanskanen, L.Niskanen, U.Tuovinen, I.Vauhkonen, E.Voutilainen |
| Kuusamo Health Center | T.Kääriäinen, E.Isopoussu |
| Kuusankoski Hospital | E.Kilkki, I.Koskinen, L.Riihelä |
| Laakso Hospital, Helsinki | T.Meriläinen, P.Poukka, R.Savolainen, N.Uhlenius |
| Lahti City Hospital | A.Mäkelä, M.Tanner |
| Lapland Central Hospital, Rovaniemi | L.Hyvärinen, K.Lampela, S.Pöykkö, T.Rompasaari, S.Severinkangas, T.Tulokas |
| Lappeenranta Health Center | P. Erola, L.Härkönen, P.Linkola, T.Pekkanen, I.Pulli, E.Repo |
| Lohja Hospital | T.Granlund, K.Hietanen, M.Porrassalmi, M.Saari, T.Salonen, M.Tiikkainen, |
| Länsi-Uusimaa Hospital, Tammisaari | I.-M.Jousmaa, J.Rinne |
| Loimaa Health Center | A.Mäkelä, P.Eloranta |
| Malmi Hospital, Helsinki | H.Lanki, S.Moilanen, M.Tilly-Kiesi |
| Mikkeli Central Hospital | A.Gynther, R.Manninen, P.Nironen, M.Salminen, T.Vänttinen |
| Mänttä Regional Hospital | I.Pirttiniemi, A-M.Hänninen |
| North Karelian Hospital, Joensuu | U-M.Henttula, P.Kekäläinen, M.Pietarinen, A.Rissanen, M.Voutilainen |
| Nurmijärvi Health Center | A.Burgos, K.Urtamo |
| Oulaskangas Hospital, Oulainen | E.Jokelainen, P-L.Jylkkä, E.Kaarlela, J.Vuolaspuro |
| Oulu Health Center | L.Hiltunen, R.Häkkinen, S.Keinänen-Kiukaanniemi |
| Oulu University Hospital | R.Ikäheimo |
| Päijät-Häme Central Hospital | H.Haapamäki, A.Helanterä, S.Hämäläinen, V.Ilvesmäki, H.Miettinen |
| Palokka Health Center | P.Sopanen, L.Welling |
| Pieksämäki Hospital | V.Sevtsenko, M.Tamminen |
| Pietarsaari Hospital | M-L.Holmbäck, B.Isomaa, L.Sarelin |
| Pori City Hospital | P.Ahonen, P.Merisalo, E.Muurinen, K.Sävelä |
| Porvoo Hospital | M.Kallio, B.Rask, S.Rämö |
| Raahe Hospital | A.Holma, M.Honkala, A.Tuomivaara, R.Vainionpää |
| Rauma Hospital | K.Laine, K.Saarinen, T.Salminen |
| Riihimäki Hospital | P.Aalto, E.Immonen, L.Juurinen |
| Salo Hospital | A.Alanko, J.Lapinleimu, P.Rautio, M.Virtanen |
| Satakunta Central Hospital, Pori | M.Asola, M.Juhola, P.Kunelius, M.-L.Lahdenmäki, P.Pääkkönen, M.Rautavirta |
| Savonlinna Central Hospital | T.Pulli, P.Sallinen, M.Taskinen, E.Tolvanen, T.Tuominen, H.Valtonen, A.Vartia, S-L.Viitanen |
| Seinäjoki Central Hospital | O.Antila, E.Korpi-Hyövälti, T.Latvala, E.Leijala, T.Leikkari, M.Punkari N.Rantamäki, H.Vähävuori |
| South Karelia Central Hospital, Lappeenranta | T.Ensala, E.Hussi, R.Härkönen, U.Nyholm, J.Toivanen |
| Tampere Health Center | A.Vaden, P.Alarotu, E.Kujansuu, H.Kirkkopelto-Jokinen, M.Helin, S.Gummerus, L.Calonius, T.Niskanen, T.Kaitala, T.Vatanen |
| Tampere University Hospital | P. Hannula, I.Ala-Houhala, R.Kannisto, T.Kuningas, P.Lampinen, M.Määttä,H.Oksala, T.Oksanen, A.Putila, H.Saha, K.Salonen, H.Tauriainen, S.Tulokas |
| Tiirismaa Health Center, Hollola | T.Kivelä, L.Petlin, L.Savolainen |
| Turku Health Center | A.Artukka, I.Hämäläinen, L.Lehtinen, E.Pyysalo, H.Virtamo, M.Viinikkala, M.Vähätalo |
| Turku University Central Hospital | K.Breitholz, R.Eskola, K.Metsärinne, U.Pietilä, P.Saarinen, R.Tuominen, S.Äyräpää |
| Vaajakoski Health Center | K.Mäkinen, P.Sopanen |
| Valkeakoski Regional Hospital | S.Ojanen, E.Valtonen, H.Ylönen, M.Rautiainen, T.Immonen |
| Vammala Regional Hospital | I.Isomäki, R.Kroneld, L.Mustaniemi, M.Tapiolinna-Mäkelä |
| Vasa Central Hospital | S.Bergkulla, U.Hautamäki, V-A.Myllyniemi, I.Rusk |
